# Social support improves nurses’ resilience: a cross-sectional study in Greece

**DOI:** 10.1101/2023.04.03.23288089

**Authors:** Petros Galanis, Aglaia Katsiroumpa, Irene Vraka, Olga Siskou, Olympia Konstantakopoulou, Theodoros Katsoulas, Parisis Gallos, Daphne Kaitelidou

## Abstract

**Background:** Since nursing job is perceived as personally and professionally demanding, internal resources such as resilience and coping skills are essential to improve nurses’ health and wellbeing and therefore work productivity and quality of patient care.

**Objective:** To assess the effect of social support on nurses’ resilience. Moreover, we investigated the impact of demographic characteristics of nurses on their resilience.

**Methods:** We conducted an on-line cross-sectional study in Greece. Data were collected during October 2022. We used the Multidimensional Scale of Perceived Social Support to measure social support, and the Brief Resilience Scale to measure resilience. We measured the following demographic characteristics of nurses: gender, age, self-perceived health status, COVID-19 diagnosis, MSc/PhD diploma, and clinical experience.

**Results:** Study population included 963 nurses with a mean age of 37.9 years. Nurses experienced moderate levels of resilience and high levels of social support. Multivariable linear regression analysis identified that increased significant others support and increased friends support were associated with increased resilience. Moreover, we found a positive relationship between age and resilience. Also, nurses with good/very good health had higher levels of resilience compared to nurses with very poor/poor/moderate health. Finally, resilience was higher among nurses with MSc/PhD diploma.

**Conclusions:** We found a positive relationship between social support and resilience among nurses. Understanding of factors that influence nurses’ resilience can add invaluable knowledge to develop and establish tailored programs. Peer support is essential to improve nurses’ resilience and promote patient healthcare.

## Introduction

A high demand job as nursing in combination with nursing shortage decrease job satisfaction and increase stress and burnout levels (Gi et al., 2011; Toh et al., 2012). Moreover, psychological issues such as depression, anxiety, stress, sleep disturbances, and burnout are prevalent among nurses especially during the COVID-19 pandemic (Al Maqbali et al., 2021; Chen & Meier, 2021; Galanis et al., 2021).

Since nursing job is perceived as personally and professionally demanding, internal resources such as resilience and coping skills are essential to improve nurses’ health and wellbeing and therefore work productivity and quality of patient care. Resilience is the ability to bounce back from adverse situations (Jackson et al., 2007). Especially, in case of nursing job resilience is related with subjective motivation to cope, self-efficacy, and emotion regulation (Stacey et al., 2019).

Formal and informal support can assist nurses to promoting resilience and coping skills (Gillman et al., 2015). Nurses should be aware of the coping strategies in order to promote their resilience (Dahl et al., 2022). Moreover, there is a positive correlation between compassion satisfaction and resilience among nurses (Unjai et al., 2022). Several strategies can be implemented in order to improve nurses’ resilience, such as improved connections within the professional team, educational programs to develop behaviors that help nurses to control or limit their stress, and tailored interventions to support nurses’ wellbeing (Gillman et al., 2015; Unjai et al., 2022).

Social support is defined as the protection and assistance given by family members, significant others and friends. A recent systematic review found a positive relationship between social support from supervisors and coworkers and burnout syndrome in nurses (Velando-Soriano et al., 2020). Several other studies confirm the positive effect of social support on nurses’ mental health such as anxiety, depression, stress, hypochondriasis, and quality of life (Chen & Meier, 2021; Li et al., 2022; Yan et al., 2022). Moreover, social support improves positive attitudes among nurses such as COVID-19 booster vaccination willingness (Galanis et al., 2022).

Until now, only three studies in Haiti, Taiwan, and China has investigated the impact of social support on nurses’ resilience (Caton, 2021; Hsieh et al., 2017; Wang et al., 2018). Therefore, no studies in developed countries have been conducted on this issue. Thus, the aim of our study was to assess the effect of social support on nurses’ resilience. Moreover, we investigated the impact of several demographic characteristics of nurses on their resilience.

## Methods

### Study design

We conducted an on-line cross-sectional study in Greece. Data were collected during October 2022. First, we used the Google forms to create an on-line version of the study questionnaire. Then, we disseminated the study questionnaire through the social media. Nurses that are working in healthcare services can participate in our study.

We measured the following demographic characteristics of nurses: gender (females or males), age (continuous variable), self-perceived health status (very poor, poor, moderate, good or very good), COVID-19 diagnosis (no or yes), MSc/PhD diploma (no or yes), and clinical experience (continuous variable).

We used the Multidimensional Scale of Perceived Social Support to measure social support that nurses receive (Zimet et al., 1988). The scale consists of 12 items and includes three factors: family support, friends support, and significant others support. Each factor takes values from 1 to 7. Higher values indicate higher levels of support. In our study, Cronbach’s alpha for the three factors ranged from 0.808 to 0.928.

Also, we measured nurses’ resilience with the Brief Resilience Scale (Smith et al., 2008). The scale includes six items and total resilience score ranges from 1 to 5. Higher values indicate higher levels of resilience. In our study, Cronbach’s alpha for the scale was 0.848.

### Ethics

We did not collect personal data of nurses. Also, we informed nurses about the aim and the design of our study and they gave their informed consent. The study protocol was approved by the Ethics Committee of Faculty of Nursing, National and Kapodistrian University of Athens (reference number; 417, 7 September 2022). Also, we applied the guidelines of the Declaration of Helsinki in our study.

### Statistical analysis

We use numbers and percentages to present categorical variables. Also, we use mean, standard deviation, median, minimum value and maximum value to present continuous variables. We calculated Cronbach’s alpha to estimate reliability of the questionnaires. Social support and demographic characteristics of nurses were the independent variables, while resilience was the dependent variable. We performed univariate and multivariable linear regression analysis to assess the impact of independent variables on nurses’ resilience. We presented unadjusted and adjusted coefficients beta, 95% confidence intervals (CI) and p-values. P-values less than 0.05 were considered as statistically significant. We used the IBM SPSS 21.0 (IBM Corp. Released 2012. IBM SPSS Statistics for Windows, Version 21.0. Armonk, NY: IBM Corp.) for the analysis.

## Results

Study population included 963 nurses with a mean age of 37.9 years (standard deviation=9.6). The majority of nurses was females (88.4%) and had a good/very good level of health (88.3%). Among nurses, 71.8% have been diagnosed with COVID-19 during the pandemic. More than half of nurses (54.6%) possessed a MSc/PhD diploma. Mean years of clinical experience was 12 (standard deviation=9.2).

Table 2 presents descriptive statistics for the scales in our study. Nurses experienced moderate levels of resilience and high levels of social support. Moreover, significant others support was higher (mean=6.15) than family support (mean=6.00) and friends support (mean=5.86).

**Table 1.** Demographic characteristics of nurses.

| Characteristics | N | % |
| --- | --- | --- |
| Gender |  |  |
| Females | 851 | 88.4 |
| Males | 112 | 11.6 |
| Age (years)<sup>a</sup> | 37.9 | 9.6 |
| Health status |  |  |
| Very poor | 26 | 2.7 |
| Poor | 16 | 1.7 |
| Moderate | 70 | 7.3 |
| Good | 580 | 60.2 |
| Very good | 271 | 28.1 |
| COVID-19 diagnosis |  |  |
| No | 272 | 28.2 |
| Yes | 691 | 71.8 |
| MSc/PhD diploma |  |  |
| No | 437 | 45.4 |
| Yes | 526 | 54.6 |
| Years of clinical experience<sup>a</sup> | 12.0 | 9.2 |

<sup>a</sup> mean, standard deviation

**Table 2.** Descriptive statistics for the scales.

| Characteristics | Mean | Standard deviation | Median | Minimum value | Maximum value |
| --- | --- | --- | --- | --- | --- |
| Resilience | 3.45 | 0.65 | 3.5 | 1 | 5 |
| Family support | 6.00 | 1.42 | 6.5 | 1 | 7 |
| Significant others support | 6.15 | 1.26 | 6.8 | 1 | 7 |
| Friends support | 5.86 | 1.38 | 6.3 | 1 | 7 |

Predictors of nurses’ resilience are shown in Table 3. Multivariable linear regression analysis identified that increased significant others support (b=0.106, 95% CI=0.059 to 0.154, p<0.001) and increased friends support (b=0.047, 95% CI=0.007 to 0.087, p=0.02) were associated with increased resilience. Moreover, we found a positive relationship between age and resilience (b=0.012, 95% CI=0.003 to 0.021, p=0.007). Also, nurses with good/very good health had higher levels of resilience compared to nurses with very poor/poor/moderate health (b=0.245, 95% CI=0.154 to 0.335, p<0.001). Finally, resilience was higher among nurses with MSc/PhD diploma (b=0.194, 95% CI=0.113 to 0.275, p<0.001).

**Table 3.** Univariate and multivariable linear regression analysis with nurses’ resilience as the dependent variable.

| Independent variables | Univariate model |  | Multivariable model |  |
| --- | --- | --- | --- | --- |
|  | Unadjusted coefficient<br>beta (95% CI) | P-value | Adjusted coefficient<br>beta (95% CI)<sup>a</sup> | P-value |
| Gender (females vs. males) | -0.099 (0.228 to 0.030) | 0.131 | -0.018 (-0.149 to 0.113) | 0.786 |
| Age (years) | 0.003 (-0.001 to 0.007) | 0.202 | 0.012 (0.003 to 0.021) | 0.007 |
| Health status (good/very good vs. very poor/poor/moderate) | 0.323 (0.234 to 0.413) | <0.001 | 0.245 (0.154 to 0.335) | <0.001 |
| COVID-19 diagnosis (yes vs. no) | -0.069 (-0.161 to 0.023) | 0.139 | -0.058 (-0.147 to 0.032) | 0.208 |
| MSc/PhD diploma (yes vs. no) | 0.171 (0.089 to 0.253) | <0.001 | 0.194 (0.113 to 0.275) | <0.001 |
| Years of clinical experience | 0.002 (-0.002 to 0.007) | 0.318 | -0.006 (-0.015 to 0.003) | 0.169 |
| Family support | 0.089 (0.060 to 0.117) | <0.001 | -0.022 (-0.060 to 0.016) | 0.252 |
| Significant others support | 0.151 (0.120 to 0.183) | <0.001 | 0.106 (0.059 to 0.154) | <0.001 |
| Friends support | 0.118 (0.089 to 0.147) | <0.001 | 0.047 (0.007 to 0.087) | 0.020 |

CI: confidence interval
<sup>a</sup> p-value for ANOVA<0.001; R<sup>2</sup> for the final multivariable model was 13.5%

## Discussion

We conducted a cross-sectional study in Greece to investigate the impact of social support on nurses’ resilience. To the best of our knowledge this is the first study on this issue in the developed countries. Additionally, we examined the impact of several demographic characteristics on nurses’ resilience.

We found that significant others support and friends support improves nurses’ resilience. Literature supports this finding since three studies in China, Haiti, and Taiwan found a positive relationship between social support and resilience (Caton, 2021; Hsieh et al., 2017; Wang et al., 2018). In particular, Wang et al. (2018) found that coworker support improves nurses’ resilience in a sample of 747 nurses in China. Also, Hsieh et al. (2017) identified a positive relationship between family and peer support and resilience, while Caton (2021) found the same relationship between social support and resilience. Social support improves the ability of nurses to better adapt to difficult conditions at the personal and professional level. Moreover, social support gives nurses the opportunity to manage effectively negative feelings that might experience during their work (Kılınç & Sis Çelik, 2021). Especially, support from significant others and friends decrease stress and anxiety that nurses experience when they work with their colleagues under the stressful environment of hospital. Moreover, older nurses can help younger colleagues to better cope with difficulties in clinical context by providing them professional help and emotional support (Welsh, 2014). Thus, peer support increases nurses’ resilience enabling them to deal with their work more effectively and better manage demanding work circumstances (Liu et al., 2018).

Among demographic characteristics, age, health status, and educational level were associated with resilience. In particular, increased age and educational level were associated with increased resilience. Also, resilience was higher among nurses with better health status. Literature supports our findings since several studies found that older age was associated with higher resilience (Cohen et al., 2014; Terrill et al., 2016; Weitzel et al., 2021). For example, a recent study in Germany found that 20% of adults over 65 years old had high resilience (Weitzel et al., 2021). Moreover, individuals whose health status was very good/excellent were more probable to exhibit higher levels of resilience compared to individuals whose health status was poor/fair/good (Hopkins et al., 2015; McKibbin et al., 2016; Soundararajan et al., 2023).

Our study had several limitations. First, we collected data through a self-administrated questionnaire. Thus, an information bias is probable in our study. Second, we used a convenience sample and therefore it is less likely to be representative of the population of nurses in Greece. Third, we performed an on-line study through social media. Therefore, selection bias is probable since nurses without accounts on social media cannot participate in our study. Fourth, we conducted a cross-sectional study and a causal relationship between social support and resilience cannot be established. Fifth, we measured several demographic characteristics of nurses but many others could be examined in future research.

In conclusion, we found a positive relationship between social support and resilience among nurses. Moreover, older nurses, those with better health, and those with higher educational level had higher levels of resilience. Since nurses work under demanding and stressful circumstances policy makers should develop a harmonious working environment in order to improve resilience and thus job productivity. Understanding of factors that influence nurses’ resilience can add invaluable knowledge to develop and establish tailored programs. Peer support is essential to improve nurses’ resilience and promote patient healthcare.

## Data Availability

All data produced in the present study are available upon reasonable request to the authors

